# The use of germicidal ultraviolet light, vaporised hydrogen peroxide and dry heat to decontaminate face masks and filtering respirators contaminated with an infectious norovirus

**DOI:** 10.1101/2020.09.04.20188250

**Authors:** Constance Wielick, Louisa F. Ludwig-Begall, Lorène Dams, Ravo M. Razafimahefa, Pierre-Francois Demeuldre, Aurore Napp, Jan Laperre, Olivier Jolois, Frédéric Farnir, Eric Haubruge, Etienne Thiry

## Abstract

In the context of the SARS-CoV-2 pandemic, reuse of surgical masks and filtering facepiece respirators has been recommended. Their reuse necessitates procedures to inactivate contaminating human respiratory and oral pathogens. We previously demonstrated decontamination of masks and respirators contaminated with an infectious SARS-CoV-2 surrogate via ultraviolet germicidal irradiation, vaporised hydrogen peroxide, and use of dry heat. Here, we show that these same methods efficiently inactivate a more resistant, non-enveloped oral virus; decontamination of infectious murine norovirus-contaminated masks and respirators reduced viral titres by over four orders of magnitude on mask or respirator coupons.

## INTRODUCTION

In the context of the ongoing severe acute respiratory syndrome coronavirus 2 (SARS-CoV-2) pandemic, the supply of personal protective equipment remains under strain and re-use of surgical face masks (masks) and filtering facepiece respirators (FFRs) has been recommended [1]. Prior decontamination is paramount to safe re-use of these items and must ideally inactivate both SARS-CoV-2 and other contaminating respiratory or oral human pathogens [2].

Human respiratory pathogens include other enveloped corona-, pneumo-, metapneumo-, paramyxo-, and orthomyxoviruses as well as non-enveloped coxsackie- and rhinoviruses; oral pathogens include astro-, picorna-, polio-, rota- and noroviruses (all non-enveloped) [3]. Enveloped viruses, surrounded by an outer lipid layer, are susceptible to harsh environmental conditions and inactivating treatments; non-enveloped viruses are known to be significantly more resistant. The small, non-enveloped human noroviruses (genus *Norovirus*, family *Caliciviridae*), recognised as the major global cause of viral gastroenteritis [4], are notorious for their tenacity in the face of decontamination [5]. The genetically and structurally similar murine norovirus (MuNoV), which replicates efficiently *in vitro*, has been identified as an appropriate surrogate virus for modelling human norovirus inactivation [6].

We previously demonstrated efficient decontamination of masks and FFRs contaminated with an infectious SARS-CoV-2 surrogate virus via ultraviolet germicidal irradiation, vaporised hydrogen peroxide, and use of dry heat [7]. In the present investigation into decontamination of MuNoV-inoculated masks and FFRs, we show that these same methods efficiently inactivate a more resistant, non-enveloped oral virus. All three methods permit demonstration of a loss of viral infectivity by more than three orders of magnitude in line with the FDA policy regarding face masks and respirators [2]. Inactivation of a norovirus, the most resistant of the respiratory and oral human viruses, can predict the inactivation of any less resistant viral mask or FFR contaminant.

## METHODS

Efficacy of three different decontamination methods in inactivating an infectious norovirus was assessed using masks and FFRs experimentally inoculated with MuNoV. Per decontamination method and mask type, one negative control mask or FFR (uncontaminated but treated), three treated masks or FFRs (MuNoV-contaminated and treated), and three positive controls (MuNoV-contaminated but untreated) were utilised. The workflow followed previously described protocols for mask and FFR inoculation, decontamination and virus elution [7].

### Virus and cells

The murine macrophage cell line RAW264.7 (ATCC TIB-71) was maintained in Dulbecco’s modified Eagle’s medium (Invitrogen) containing 10% heat inactivated foetal calf serum (FCS) (BioWhittaker), 2% of an association of penicillin (5000 SI units/ml) and streptomycin (5 mg/ml) (PS, Invitrogen) and 1% 1 M HEPES buffer (pH 7.6) (Invitrogen) at 37 °C with 5% CO_2_.

Stocks of MuNoV isolate MNV-1.CW1 were produced by infection of RAW264.7 cells at a multiplicity of infection of 0.05. Two days post-infection, cells and supernatant were harvested and clarified by centrifugation for 20 minutes at 1000 x g after three freeze/thaw cycles (– 80°C/37°C).

Titres were determined via the tissue culture infective dose (TCID_50_) method; RAW 264.7 cells were seeded in 96-well plates, infected with 10-fold serial dilutions of MuNoV, incubated for three days at 37 °C with 5% CO_2_, and finally stained with 0.2% crystal violet for 30 minutes. Titres, expressed as TCID_50_/ml, were calculated according to the Reed and Muench transformation [8]. A virus stock with a titre of 10^7.06^ TCID_50_/ml was used in subsequent steps.

### Surgical masks and filtering facepiece respirators

All masks and FFRs, verified to be from the same respective manufacturing lot, were supplied by the Department of the Hospital Pharmacy, University Hospital Centre of Liege (Sart-Tilman). Manufacturers (and models): KN95 FFR - Guangzhou Sunjoy Auto Supplies CO. LTD, Guangdong, China (2020 N°26202002240270); surgical mask (Type II) - Hangzhou Sunten Textile Co., LTD, Hangzhou, China (SuninCare™, Protect Plus).

### Murine norovirus inoculation of surgical masks and filtering facepiece respirators, decontamination, elution and quantification

Per treated or control mask or FFR, 100 µl of undiluted viral suspension were injected under the first outer layer at the centre of each of three square coupons (34 mm x 34 mm). In addition to inoculation of the *de facto* masks or FFRs, 100 µl of viral suspension was pipetted onto one elastic strap per contaminated mask or FFR. Masks and FFRs were allowed to dry for 20 minutes at room temperature before decontamination via UV irradiation, vaporised H_2_0_2_, and dry heat.

Masks and FFRs were individually UV-irradiated for 2 minutes (2.6J/cm^2^ fluence per mask), using a LS-AT-M1 (LASEA Company, Sart Tilman, Belgium). Vaporous hydrogen peroxide (VHP) treatment was performed with the V-PRO Max Sterilizer (Steris, Mentor, OH) which uses 59% liquid H_2_O_2_ to generate hydrogen peroxide vapor. A 28-minute non lumen cycle was used, consisting of 2 min 40 sec conditioning (5 g/min), 19 min 47 sec decontamination (2.2 g/min) and 7 min 46 sec aeration (750 ppm peak VHP concentration). Dry heat decontamination was performed at temperatures of 102°C (± 4°C) for 60 min (± 15 min) in an electrically heated vessel (M-Steryl, AMB Ecosteryl Company, Mons, Belgium).

Upon completion of the decontamination protocols, MuNoV was eluted from three excised coupons and one severed elastic strap per mask or FFR into 4 mL elution medium (Eagle’s DMEM (Sigma)) supplemented with 2 % of an association of penicillin (5000 SI units/mL) and streptomycin (5 mg/mL) (PS, Sigma) and, for elution from VHP-treated materials, 20% FCS and 0.1% β-mercaptoethanol) via 1 minute (coupons) or 20 minute vortex (straps) at maximum speed (2500 rounds per minute; VWR VX-2500 Multi-Tube Vortexer).

Titres of infectious MuNoV recovered from individual coupons and straps were determined via TCID_50_ assay. Back titrations of inoculum stocks were performed in parallel to each series of decontamination experiments.

### Data analysis and statistics

All statistical analyses were performed using SAS® software 9.3 (SAS/ETS 12.1 - SAS STAT 12.1). Linear mixed models were studied using the MIXED procedure; in addition, TOBIT models were implemented using the qualitative and limited dependent variable model (QLIM) procedure. All p-values reported using the QLIM procedure were obtained using Wald tests.

## RESULTS

Back titrations of virus inoculums performed in parallel to each series of experiments confirmed MuNoV inoculum titres to be within a range of 3.55×10^7^ to 6.31×10^7^ TCID_50_/mL for all experiments.

The cell culture limit of detection (LOD) was 0.8 log10 TCID_50_/ml for all analyses except those concerning H_2_O_2_-treated mask- or FFR straps (2.8 log10 TCID_50_/ml) and UV-treated FFR straps (1.8 log10 TCID_50_/ml).

High levels of infectious virus were recovered from MuNoV-inoculated, untreated coupons of all masks and FFRs, with mean overall recovery values of 4.94 (±0.55 standard deviation (SD)) log10 TCID_50_/mL. Mean strap recovery values were similar between experiments, however they were lower than coupon recovery values (notable exception: elution from untreated masks), with mean values of 4.11 (±0.77 SD) log10 TCID_50_/mL (Figure 1).

**Figure 1.**
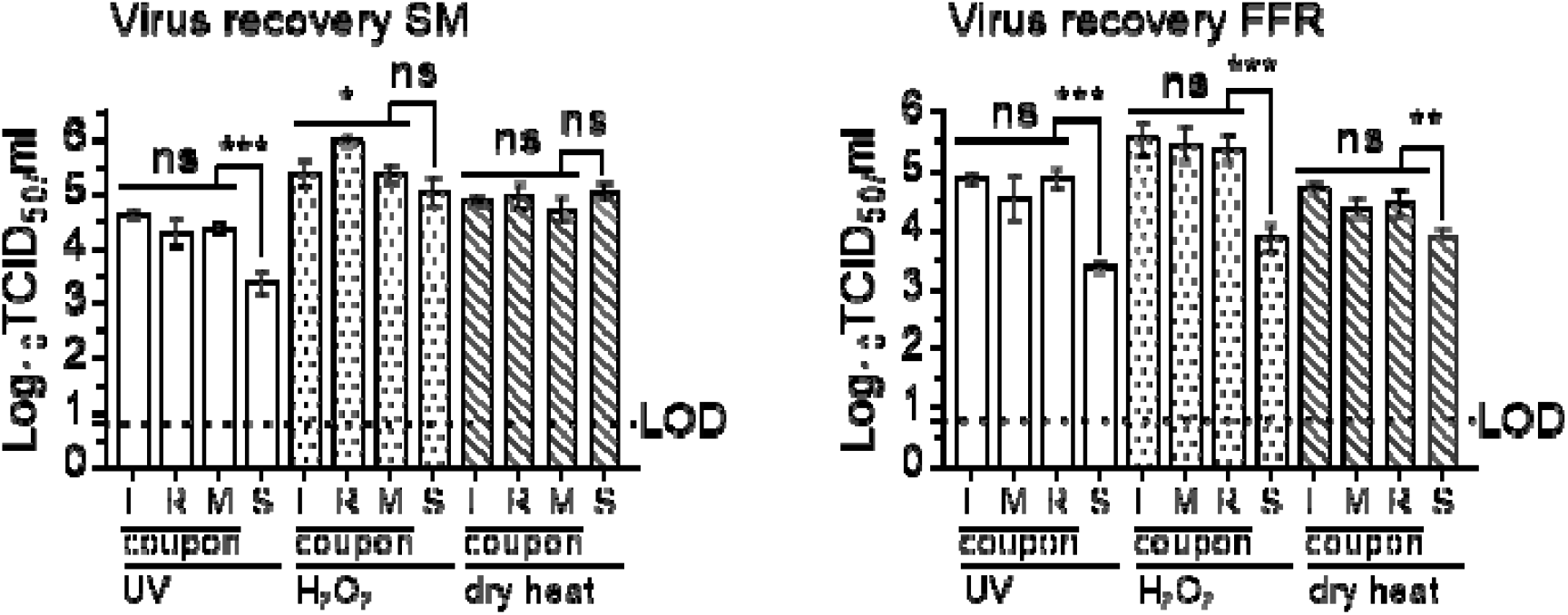
Recovery of MuNoV after elution from inoculated, untreated surgical masks and filtering facepiece respirators. Recovery of infectious murine norovirus (MuNoV) from inoculated untreated surgical masks (SM) and filtering facepiece respirators (FFR) was analysed in RAW 264.7 cells. The cell culture limit of detection (LOD) was 0.8 log_10_ TCID_50_/mL. Similar levels of virus recovery were detected for left, right and middle (L, R, M) coupons of masks and respirators; recovery efficacy of infectious virus from straps (S) deviated significantly in all analyses from the mean of all coupons (with the exception of extraction from SM straps in the H_2_O_2_ assay). Statistical analyses were performed using SAS® software 9.3 (SAS/ETS 12.1 - SAS STAT 12.1). Linear mixed models were studied using the MIXED procedure; in addition, TOBIT models were implemented using the qualitative and limited dependent variable model (QLIM) procedure. All p-values reported using the QLIM procedure were obtained using Wald tests. P-values were computed to calculate differences between individual coupon values and differences between mean values of all coupons and straps: ***P<0.001, **P<0.01, *P<0.05, and ns is P≥0.05.

**Figure 2.**
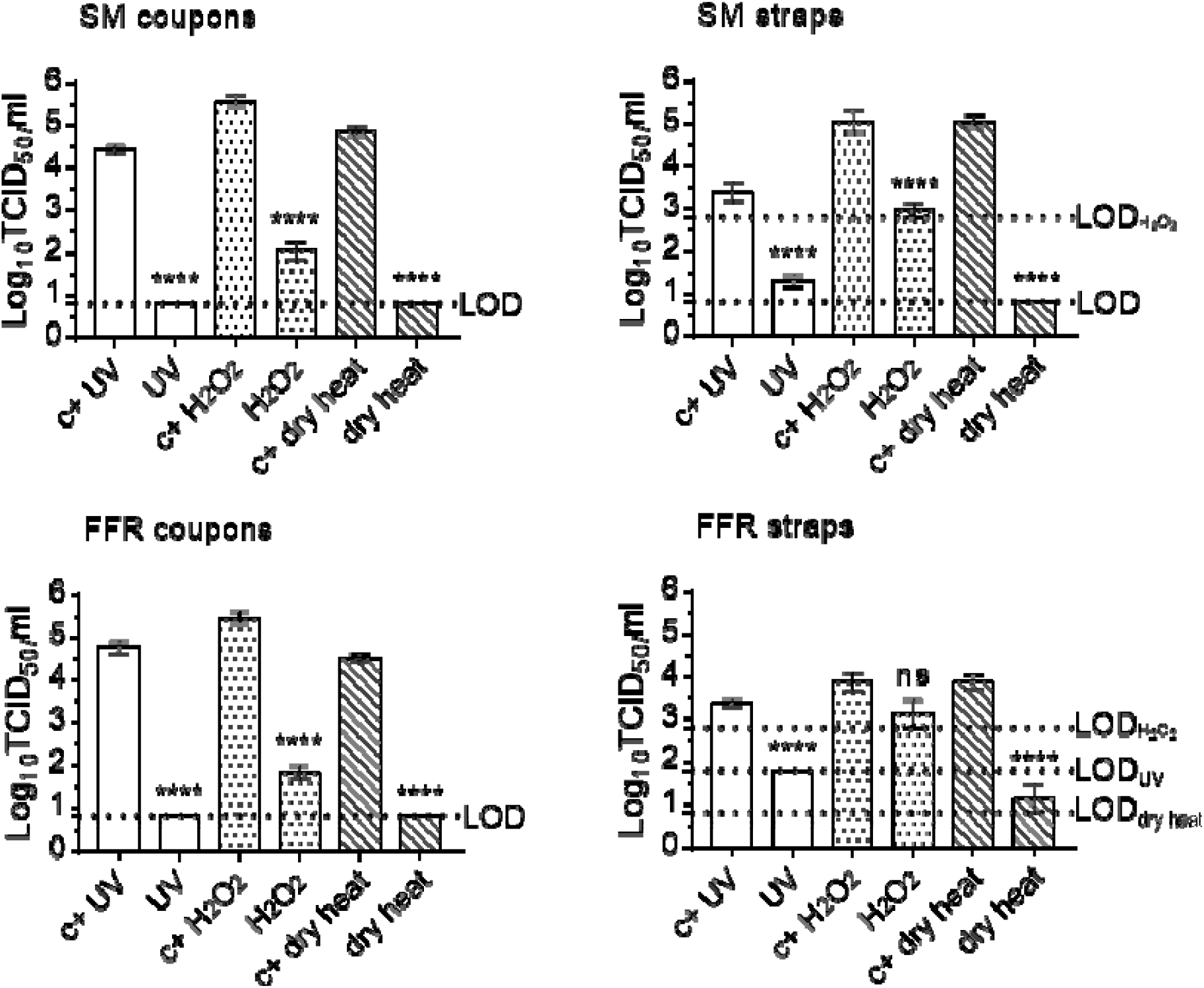
Effect of three decontaminating treatments on MuNoV-inoculated surgical mask- and filtering facepiece respirator coupons and straps. The infectivity of murine norovirus (MuNoV) recovered from surgical masks (SM) and filtering facepiece respirators (FFR) decontaminated via exposure to ultraviolet light (UV), vaporised hydrogen peroxide (H_2_O_2_), or dry heat treatment was analysed in RAW 264.7 cells. The cell culture limit of detection (LOD) was 0.8 log_10_ TCID_50_/ml for all analyses except those concerning H_2_O_2_-treated SM or FFR straps (1.8 and 2.8 log_10_ TCID_50_/ml, respectively) and UV-treated FFR straps (1.8 log_10_ TCID_50_/ml). Per decontamination method, nine MuNoV-inoculated, decontaminated coupons (n=9) and three inoculated, decontaminated straps (n=3) were analysed in parallel to inoculated, untreated, positive control coupons (n=9) and straps (n=3). Statistical analyses were performed using SAS® software 9.3 (SAS/ETS 12.1 - SAS STAT 12.1). Linear mixed models were studied using the MIXED procedure; in addition, TOBIT models were implemented using the qualitative and limited dependent variable model (QLIM) procedure. All p-values reported using the QLIM procedure were obtained using Wald tests; ****P<0.0001; ***P<0.001, **P<0.01, *P<0.05, and ns is P≥0.05.

Following mask UV irradiation and dry heat treatment, titres for virus recovered from coupons remained below the assay LOD, showing total loss of infectivity of around four orders of magnitude (3.64 (±0.28 SD) log10 TCID_50_/mL and 4.06 (±0.30 SD) log10 TCID_50_/mL, respectively), while titres of virus recovered from H_2_O_2_-vaporised coupons indicated a loss of infectivity of four orders of magnitude (4.06 (±0.30 SD) log10 TCID_50_/mL). Titres of virus recovered from treated mask straps were reduced by two orders of magnitude post UV irradiation and H_2_O_2_-treatment (2.06 (±0.29 SD) log10 TCID_50_/mL, and 2.08 (±0.38 SD) log10 TCID_50_/mL, respectively) and by four orders of magnitude post heat-treatment (4.25 (±0.25 SD) log10 TCID_50_/mL (below LOD)).

Decontamination followed a similar pattern of viral inactivation for UV-treated FFR coupons, reducing viral titres by around four orders of magnitude (3.97 (±0.40 SD) log10 TCID_50_/mL). Following vaporised H_2_O_2_- and dry heat-treatment, titres for virus recovered from coupons showed a loss of infectivity of three orders of magnitude (3.72 (±0.29 SD) log10 TCID_50_/mL and 3.64 (±0.66 SD) log10 TCID_50_/mL, respectively). UV-, H_2_O_2_- and heat-treatment of FFR straps reduced infectivity by to a lesser degree (1.58 (±0.14 SD) log10 TCID_50_/mL, from 3.38 (±0.14 SD) log10 TCID_50_/mL to below the LOD, 0.75 (±0.90 SD) log10 TCID_50_/mL and 2.75 (±0.50 SD) TCID_50_/mL, respectively).

## CONCLUSIONS

This is, to our knowledge, the first description of stable disinfection of surgical masks and FFRs contaminated with an infectious norovirus using UV irradiation, vaporised H_2_O_2_, and dry heat treatment.

Here we demonstrate successful recovery of high quantities of infectious MuNoV from inoculated, otherwise untreated masks and FFR coupons. Three decontamination methods, chemical vaporised H_2_O_2_ and physical inactivation via UV irradiation and dry heat treatment, successfully reduced infectious loads of MuNoV inoculated under the outer surface layer of mask and FFR coupons by more than three orders of magnitude. Since carrier surfaces likely influence decontamination efficacy, we examined viral inactivation not only on the *de facto* FFRs or masks, but also on their elastic straps that may become equally contaminated. We compared titres of infectious virus recovered from inoculated, untreated mask or FFR straps and those inoculated and subsequently decontaminated. While all three decontamination methods were successfully validated as they reduced viral loads by at least more than three orders of magnitude, the elevated LOD of UV-treated FFR straps and H_2_O_2_-vaporised mask- and FFR straps prevented detection of higher infectivity losses. Further studies are planned to elucidate these effects, which may potentially be associated either to inherent virucidal properties of or poor elution from the elastic materials.

In conclusion, we describe successful validation of three decontamination methods, UV irradiation, vaporised H_2_O_2_, and dry heat treatment, in inactivating an infectious non-enveloped virus in line with the FDA policy regarding face masks and FFRs. The MuNoV surrogate supplements existing data regarding decontamination of surgical masks and FFRs, and both it and the different decontamination methods tested, are easily adaptable to other FFR and mask types, presenting a useful conservative model for stable validation of non-enveloped virus decontamination.

## Data Availability

The authors confirm that the data supporting the findings of this study are available within the article.

## ACKNOWLEDGEMENTS

The authors express their sincere gratitude to Amélie Matton and Frédéric de Meulemeester (AMB Ecosteryl, Mons, Belgium), Axel Kupisiewicz (LASEA, Sart-Tilman, Belgium), Pierre Leonard (Solwalfin, Belgium) for suggestions and technical and administrative support.

## CONFLICTS OF INTEREST STATEMENT

The authors have no conflicts of interest to disclose.

## FUNDING SOURCE

This work was supported by grants from the Walloon Region, Belgium (Project 2010053 -2020-“MASK - Decontamination and reuse of surgical masks and filtering facepiece respirators”) and the ULiège Fonds Spéciaux pour la Recherche 2020.

## Non-standard abbreviations

FFR: filtering facepiece respirator
SM: surgical mask
MuNoV: murine norovirus
SARS-CoV-2: severe acute respiratory syndrome coronavirus 2

## Notes

### Competing Interest Statement

The authors have declared no competing interest.

### Author Declarations

No IRB/oversight body approval was needed. The study did not involve human subjects and is thus exempt.

